# A logistic model of CoV-2 propagation

**DOI:** 10.1101/2020.07.20.20157826

**Authors:** Robert F. Weiss

## Abstract

We describe an elemental logistic model for the propagation of CoV-2 in a community and illustrate the sensitivity of the model to key parameters such as **R**_**0**_, the initial rate of infections per infected person, and **A**_**0**_, the fraction of infected people developing neutralizing antibodies. We demonstrate the importance of the **duration** of immunity in the population, the development of waves of new cases of infection, and the effect of premature opening of local economies.

## Introduction

The CoV-2 pandemic has been controlled in countries with strict enforcement of quarantine, but appears to be out of control in the US, Brazil, Russia and India [1, 2]. Even in US states that initially avoided outbreaks, or successfully mitigated and then diminished the disease, there is great concern over the possibility of a “second wave’’ of infection during the flu season at the end of the year, or even beyond 2020 if an effective vaccine is not available.

Predictive epidemiological simulations, as opposed to retrospective data fits, are useful for both healthcare planning and policy decisions at the state and federal levels. Simulations are necessarily approximations of the dynamics of disease spread and control, but must still bear a reasonable resemblance to prior data and new case time series under different scenarios. They must also be capable of determining the effectiveness of quarantine, the development of antibodies in the infected population, and the duration of either disease-induced or vaccine-induced immunity. Finally, a simulation model should provide some insight into the probability of achieving “herd immunity”-- when a population is allowed some exposure to the virus in order to build immunity among the general population.

In this study, we describe an elemental “logistic” model [3] and the assumptions on which it is based. This will be followed by illustrations of the sensitivity of the model to both the assumptions and key parameters, such as R_**0**_, the commonly cited infection rate parameter; i.e., the rate of viral spread per infected person.

### Simulation Model

Our model is grounded in the observation that the rate of spread, or mitigation, of CoV-2 is determined not only by R_**0**_, but also by the time-dependent fractions of both the infected and uninfected populations in a defined environment. A “defined environment” is a neighborhood, town, or small city, in which “community spread” occurs. We are not attempting to simulate the dynamics of infection in major cities or entire states, much less in the entire country. Instead, we assert that community spread is primarily a “microscale” phenomenon that is a function of population density. In densely occupied apartments and buildings, nursing homes and community gathering places-- -also in bars, gyms and other small businesses—as quarantine guidelines are relaxed, community spread increases.

What is reported as new cases in cities and states is a “macroscale” metric, the sum of a large number of “microscale” events that can occur simultaneously or spread over many weeks, as well as in mass indoor gatherings for political events. Of course, there will be a continuous interaction between these two scales, as between neighborhoods and boroughs of a city, and between separate cities and states on multi-month time scales.

Because CoV-2 is both highly infectious and asymptomatic for weeks, there is a critical time delay between infection, spread, and quarantine/hospitalization. At the “microscale”, a small number of infections can grow rapidly without significantly depleting the broader, uninfected population. As the infected population grows, it locally encounters a relatively smaller fraction that is *uninfected*, arresting the rate of spread, followed by a “flattening of the curve” and eventual decline in new cases.

The infected population is then either totally or partially immune, or it is symptomatic and withdraws from the population via quarantine, hospitalization, or death. If long-duration immunity is achieved in most of the population, it will present fewer and fewer opportunities for infection and eventually eliminate the disease. This is what is meant by “herd immunity”.

However, if immunity is of limited duration, the infection rate will reach a minimum and then increase in a “second wave”. In the limit of negligible immunity in the population, repeated infections can occur on a weekly basis: the newly infected, initially asymptomatic, population will always encounter a new cohort of the vulnerable population and repeated cycles of new case growth will occur on shorter time scales. We will examine each of these scenarios.

Infection/symptomatic time lags and “feedback” from the immune population can greatly amplify the already exponential growth of infections, but they also suggest a model that predicts new cases at discrete time steps that are approximately the difference between the infection event and the appearance of symptoms. A simple time step approximation of the well-known *logistic equation* [3] readily simulates both short and long -term cycles of infection and immunity. It takes the following form:

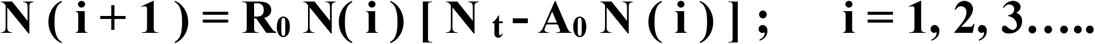

Here N (i +1) is the per cent of new cases in the population at time step (i+1), N(i) is the previously infected population at step (i), N _t_ is the time-dependent uninfected population, and [N _t_ - A_**0**_ N (i)] is the vulnerable population in the following time period, where A_**0**_ is the fraction of infected persons who have developed effective CoV-2 antibodies.

At early times and small infected populations, the infection rate is driven by N (i); as the infected population increases, with immunity determined by A_**0**_, the uninfected population decreases, thereby limiting further growth in new cases. As the new case fraction diminishes further over the following weeks, the uninfected population in the control space either gradually increases or, by relaxation of quarantine policies, is immediately expanded.

It is noted again that N (i) is the infected *fraction* of the total population at a discrete time step and is completely general; the actual infected population is determined by the total population in the control space being monitored. An important question is whether or not “herd immunity” can be achieved under varying durations of immunity and realistic quarantine policies, and at what *fraction* of the population could this occur.

The logistic equation output is easily computed with an Excel spreadsheet; the resulting weekly new case population is plotted against time steps 1-25 in the illustrations that follow. The relation between these steps and physical time is determined by the number of weeks between infection and the appearance of symptoms. This time scale is currently assumed to be between one and two weeks; we will initially assume the former. When more complete data is available, the physical time scale can be adjusted.

We can estimate the range of values for R_**0**_ by recognizing that, R_**0**_ is the product of three factors: the contact rate per infected person, the infection rate per contacted person, and the elapsed time between the contact and the appearance of symptoms leading to quarantine or hospitalization. Assuming the number of contacts in the elapsed time is between one and five, and the infection rate is between 0.5 and 1.0, their product ranges from 0.5 to 5. Obviously, at the lower end of this range there is no pandemic, while at the upper end it is catastrophic.

As we demonstrate, values of R_**0**_ between 1.0 and 2.0 produce infected population fractions that can be controlled in a single “wave”, while those between 2.0 and 4.0 can lead to multiple waves and even “chaotic” behavior, depending on the durability of or vaccination.

### Parametric Results

Three scenarios will be explored: (1) no enduring immunity: N _t_ =1.0; (2) “permanent” immunity: N _t_ < 1.0 and decreases to zero; (3) limited duration immunity: N _t_ < 1.0 but never decreases to zero. We first examine the model’s behavior under scenario (1) around the critical value of R_**0**_ = 1.0 with A_**0**_ = 1.0; i.e., all those infected achieve immunity in only a week or two. In Fig. 1, new weekly cases for values of R_**0**_ = 0.9 and 1.1 are illustrated with an initial population of 100 cases per 100,000 population; i.e., N (0) = 10 ^**- 3**^. We will assume this initial condition in other examples, as it is a common metric used to monitor the spread of the pandemic. This initial fraction of the population only determines the week in which the simulation begins. Absolute populations for any control space can obviously be derived by multiplying by the total population in the community. The ordinate scale in all of the following figures is the weekly new case *fraction* of the total population.

**Fig. 1.**
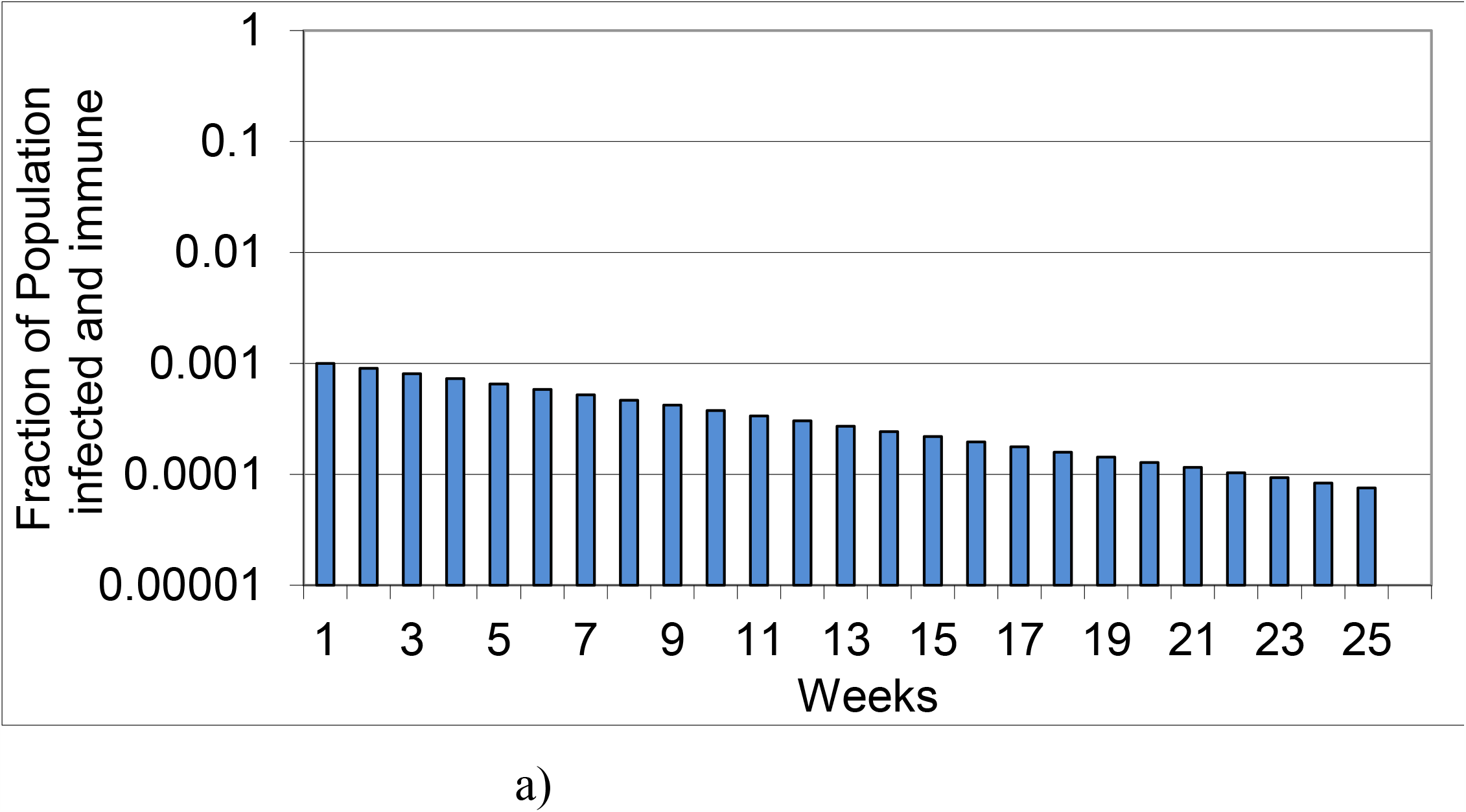
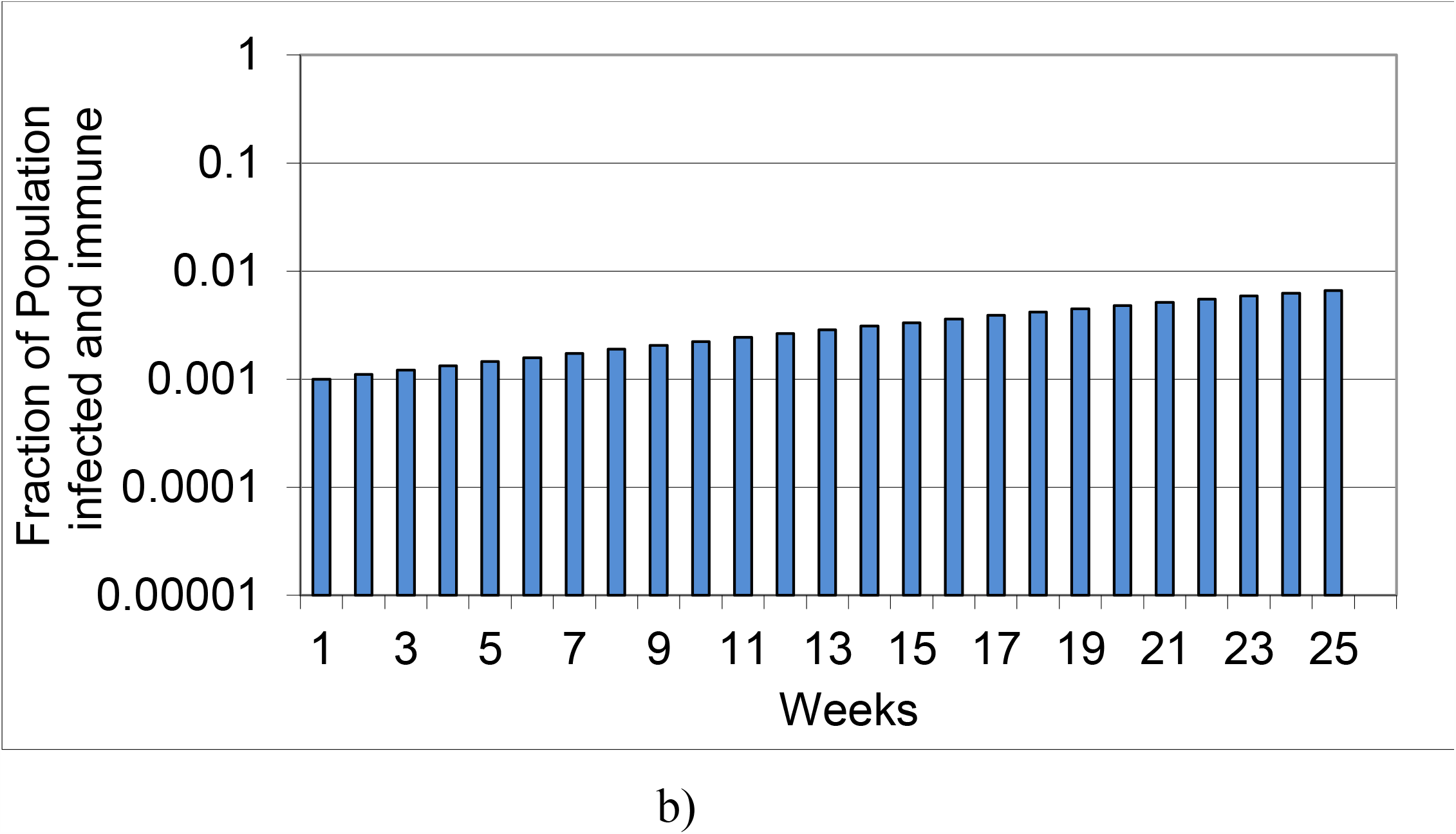
Simulations of infected population fraction for N (0) = 10 ^**– 3**^; R_**0**_ = a) 0.9; b) 1.1; A = 1.0.

We note the sensitivity to R_**0**_, but also the slow decay or growth of even initially small fractional populations. In each of these examples, it was assumed that the infected population achieved *no immunity* for the *entire* time of the simulation; scenario (1).

In Fig. 2, scenario (2) is assumed: each set of new cases of infection results in *immunity* for the *entire* time of the simulation. It also shows the effect of a larger value of R_0_ = 1.5.

**Fig. 2.**
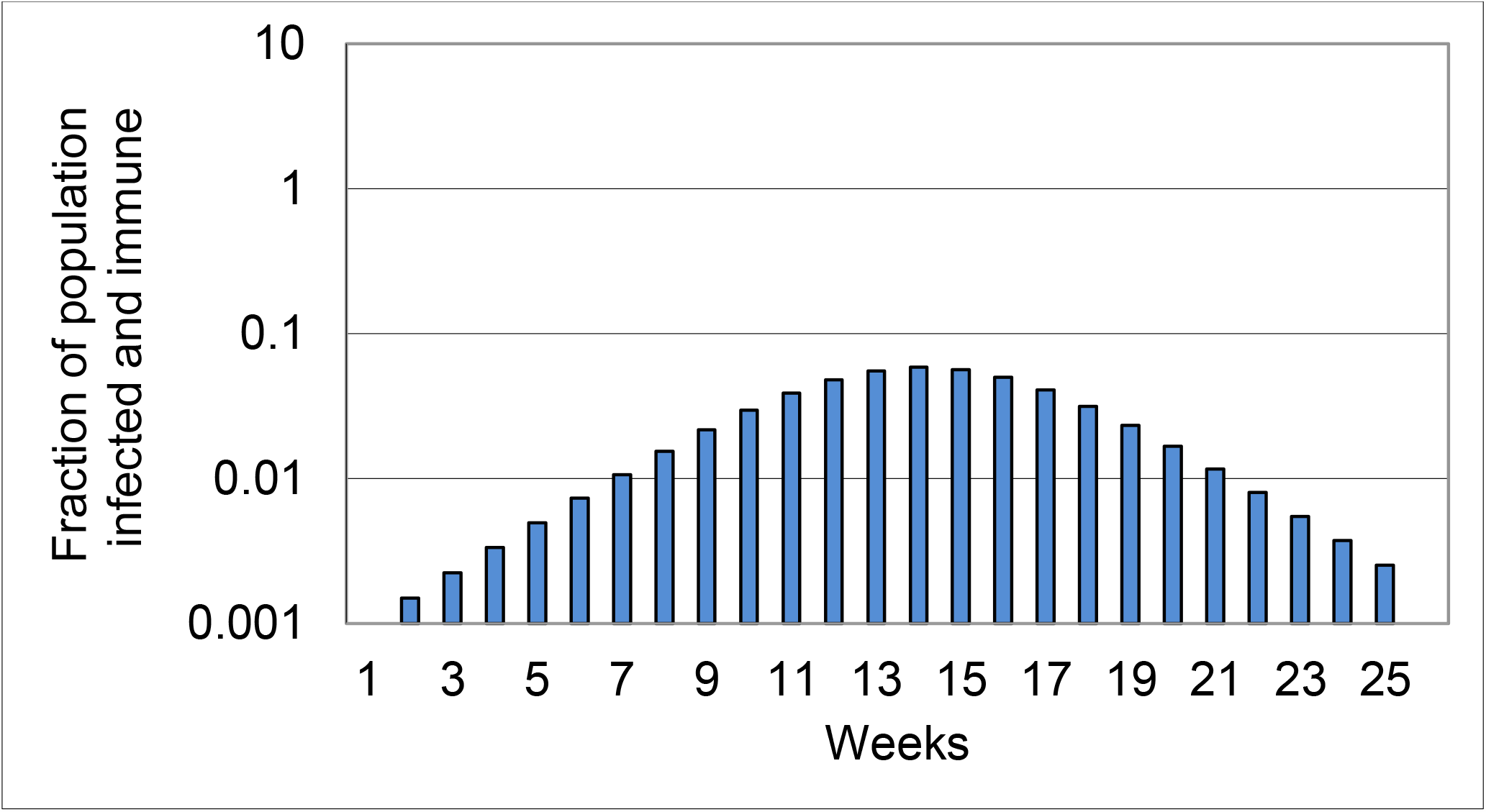
Simulation of weekly new case infections that confer “permanent” immunity and eradication of the virus: R_0_= 1.5, A = 1

In this scenario, following a peak of approximately 80 times the initial infection fraction, weekly new case counts decline by a factor of 40, to only a few cases by week 25, as expected.

We next consider scenario (3), the effect of limited duration immunity; i.e., only the *most recent six weeks of infections* in the population conferred immunity. This simulation removes the infected population from the initial total population for only the preceding six weeks, with N_t_ decreasing each of these weeks.

Here, after weekly new cases peak in week 15, they plateau in week 22. This lack of “permanent” immunity is then manifested by a resurgence of new cases in week 25. Moreover, under political pressure to re-open the economy, many states have phased in access to businesses and places of entertainment. Early openings have seen an explosion of new cases as R_0_ has obviously increased.

In Fig. 4, we simulate the effects of doubling contacts per person per week, thereby doubling R_0_from 2.0 to 4.0 at week 15 in the same two examples: permanent” immunity and “limited duration” immunity. In the former scenario, the downward trajectory of weekly new cases is disrupted for five weeks before continuing. In the latter example, there is an immediate increase in cases, and the appearance of second and third waves. Moreover, weekly new cases exceed 200 times their initial value, and continue to oscillate between 200 and 80. This behavior is a manifestation of the quasi-chaotic behavior of the basic logistic equation (**N** _**t**_ = 1) for values of R_0_ > 3.5 [3].

**Fig. 3.**
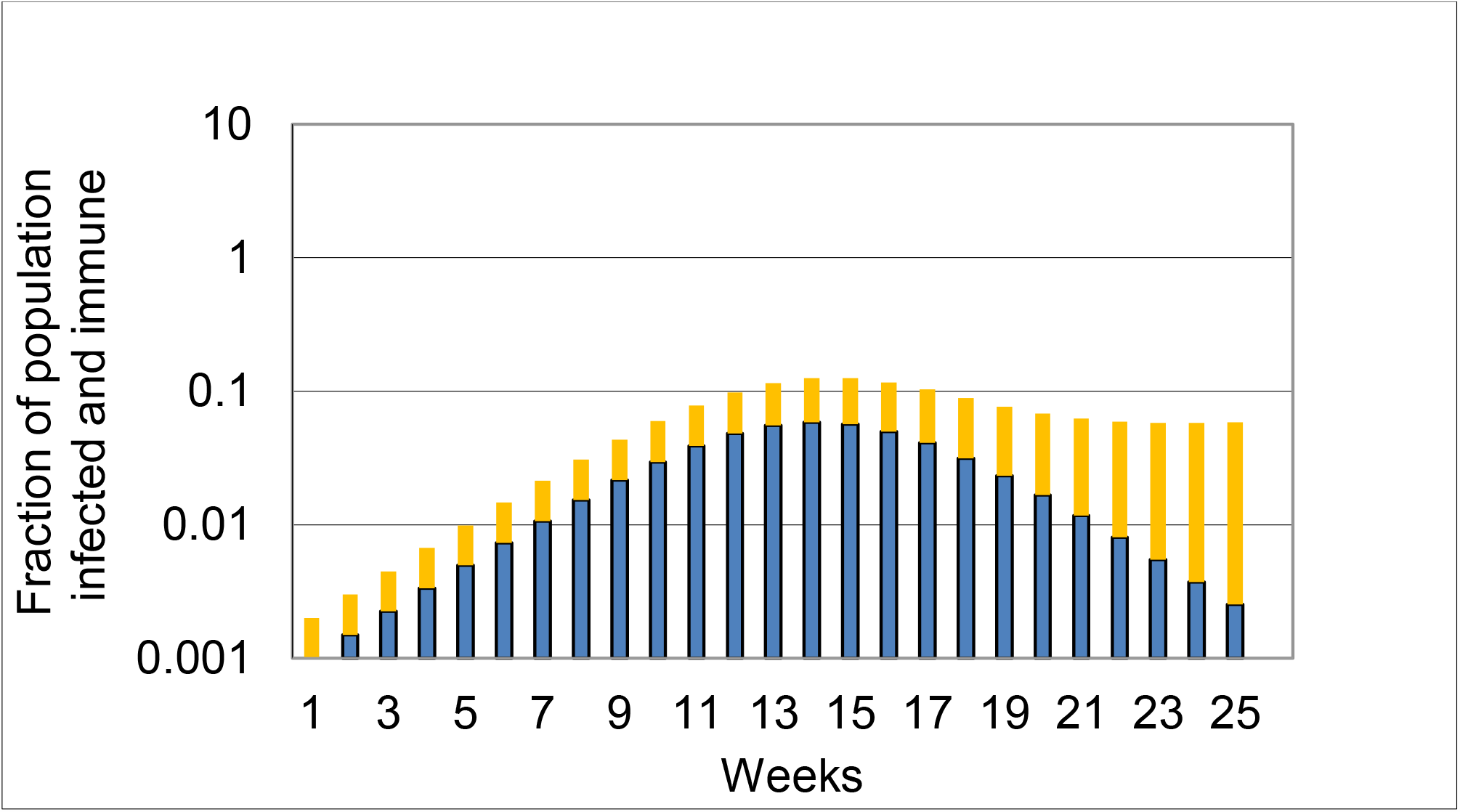
Permanent immunity (blue) compared to limited duration immunity (orange) for R_0_= 1.5, A = 1.

**Fig. 4.**
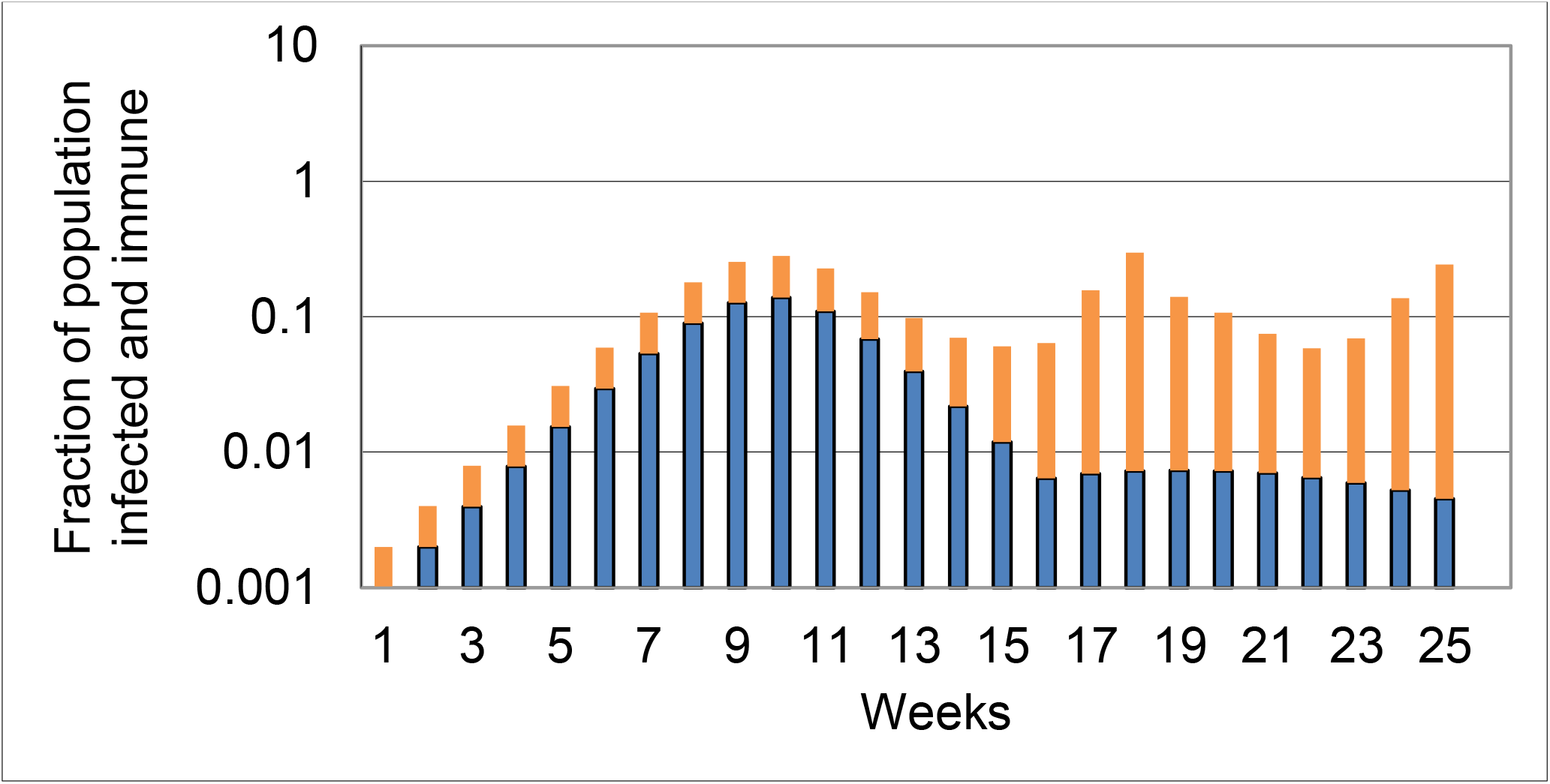
Simulation of “opening” at week 15: permanent (blue) vs limited duration immunity (orange).

Finally, in Fig. 5 we illustrate the effect of incomplete development of neutralizing antibodies, resulting in re-infection in the general population [4].

**Fig. 5.**
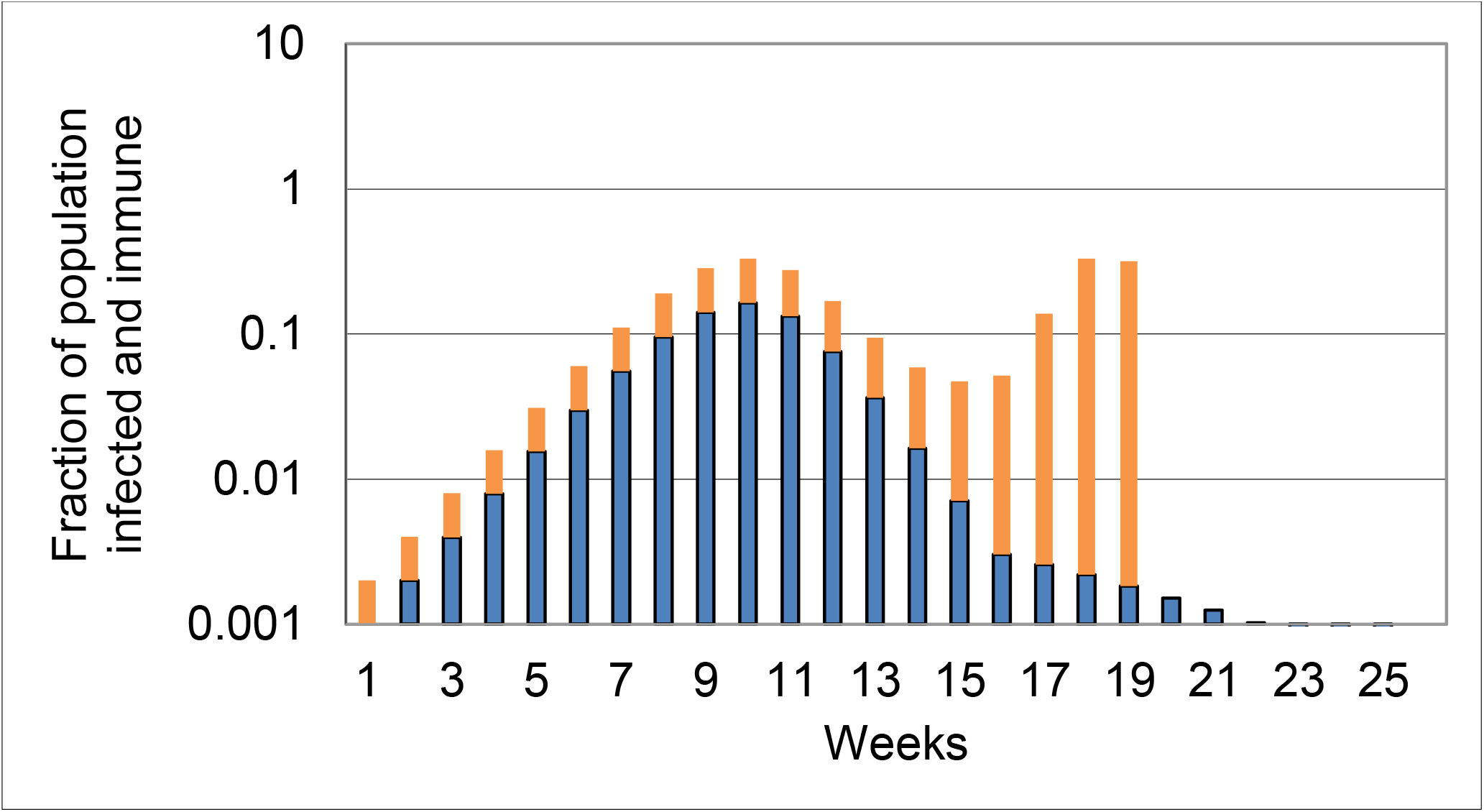
Incomplete development of neutralizing antibodies in 50% of the infected population; A_**0**_ = 0.5 (Fig.4 scenarios).

The somewhat counter-intuitive results in Fig. 5 are illustrative of the effects of “herd immunity”; a reduction in acquired immunity, combined with limited duration of immunity (orange plot), cause a spike in new cases followed by a collapse of the uninfected population. In this example, 66% of the population is infected before the collapse occurs. In the “permanent” immunity scenario, there is barely any effect of a 50% reduction in acquired immunity and the total declines steadily, even in the event of a relaxation of the quarantine.

Finally, cities such as Boston and St.Louis [5] have successfully mitigated CoV-2 outbreaks by relatively rapid reduction of R_0_, as demonstrated in Fig. 6.

**Fig. 6.**
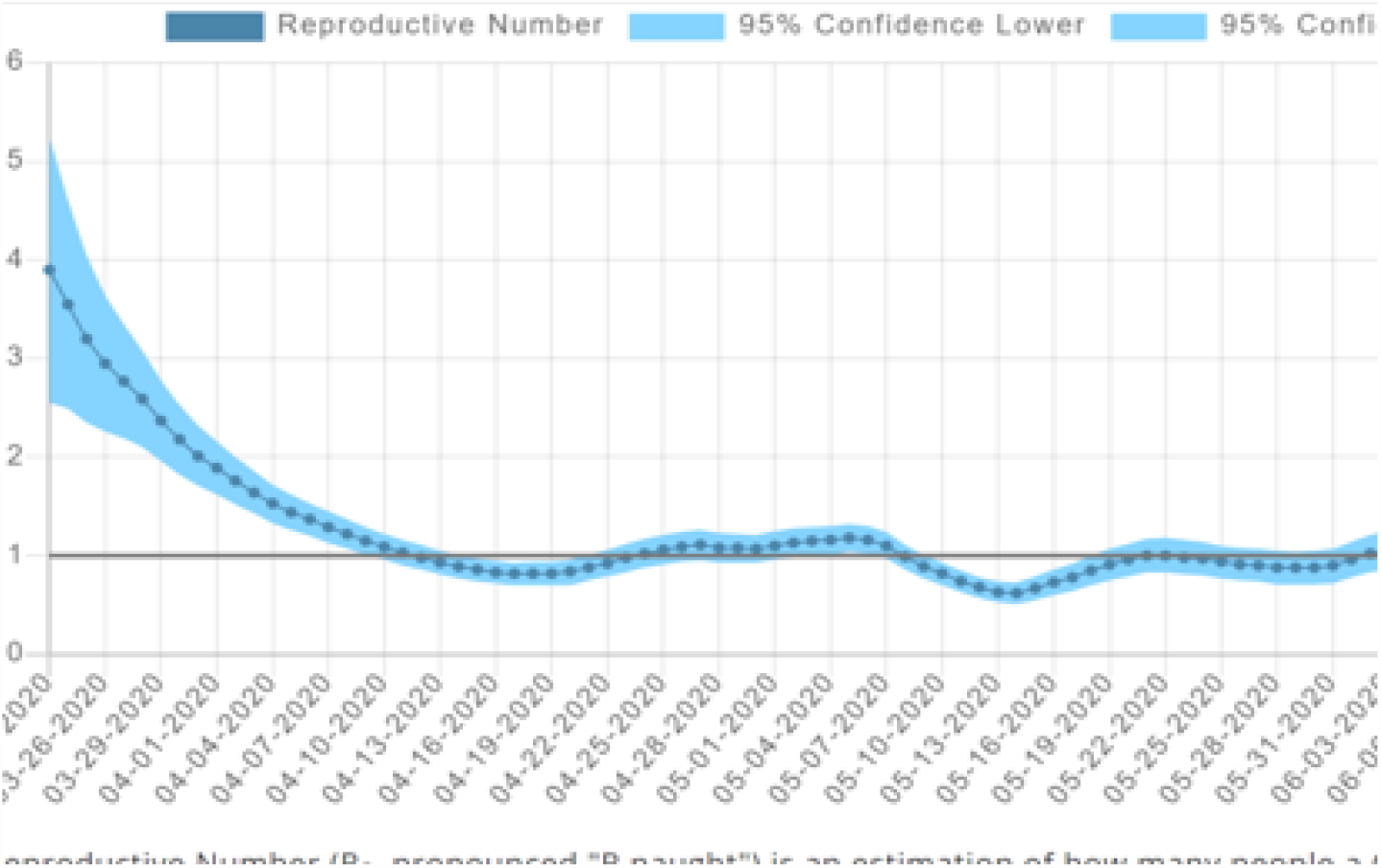
Derived CoV-2 infection rate history for St. Louis, MO [5].

Rapid infection rates, with R_0_ in the range of 2.5 to 5.0, were derived from the early data, consistent with our estimates. Even where the infection rate has declined to 1.0, it continues to oscillate above and below this critical value. As illustrated in Fig. 1, it takes many weeks for the disease to be under control, and the 7-day new case average was almost constant;

### Summary

In this study, we have applied the classic logistic equation in difference form to simulate the behavior of CoV-2 community spread and critical parameters that drive the spread. We illustrate its sensitivity to the infection rate R_0_ and the neutralizing antibody fraction A_**0**_. ***An important prediction of the model is its sensitivity to the duration of immunity, six weeks generating repeated waves of infection***, in contrast to the steady decline of new cases under a “permanent” immunity scenario. We have also examined the dramatic effect of a relaxation of mandatory quarantines by positing just a doubling of the contact rate.

Our simulated results show a rapid return to earlier levels of new cases in the limited duration immunity scenario, and a lengthy delay of disease eradication even with the development of “permanent” immunity in the population. According to our model, “herd immunity” develops in the former scenario only when weekly cases reach 66% of the population, while 20% is required in the latter scenario.

Our model has not, and cannot, address the detailed modes of disease transmission, but can suggest the trends of disease spread with and without quarantine and contact tracing to prevent future outbreaks. The importance of limited duration immunity is clearly indicated. Finally, our model suggests that herd immunity may never occur. Detailed analysis of new case data in specific cities, particularly those that have recently invoked a rapid “opening” policy, might provide useful policy guidance to federal and state officials.

## Data Availability

All data presented in the manuscript is available immediately

## Acknowledgements

*The author acknowledges the contributions of David H. Alpers, MD, David E. Wolf, PhD, and John F. Cronin, as well as comments from Steven G. Krantz, PhD and Arni Srinivasa Rao, PhD*.

## Footnotes

* It is not a “safe bet” to rely on immunity to Covid-19 as a strategy for coping with the pandemic, one expert has warned, adding that herd immunity strategies were “probably never going to work.” Speaking to CNBC’s “Squawk Box Europe” on Monday, Danny Altmann, professor of immunology at Imperial College London, said that in towns and cities where there had been coronavirus infections, only 10% to 15% of the population was likely to be immune.”And immunity to this thing looks rather fragile — it looks like some people might have antibodies for a few months and then it might wane, so it’s not looking like a safe bet,” he said. “It’s a very deceitful virus and immunity to it is very confusing and rather short lived.” He also raised questions about the likely success of so-called herd immunity — when a population is allowed some exposure to the virus in order to build immunity among the general

